# COVID-19 and non-Hodgkin’s lymphoma: a common susceptibility pattern?

**DOI:** 10.1101/2022.11.01.22281794

**Authors:** De Matteis Sara, Cosetta Minelli, Giorgio Broccia, Paolo Vineis, Pierluigi Cocco

## Abstract

**Objectiv:** To explore the link between COVID-19 incidence, socio-economic covariates, and NHL incidence.

**Design:** Ecological study design.

**Setting:** Sardinia, Italy.

**Participants:** We used official reports on the total cases of COVID-19 in 2020, published data on NHL incidence, and socio-economic indicators by administrative unit, covering the whole regional population.

**Main outcomes and measures:** We used multivariable regression analysis to explore the association between the natural logarithm (ln) of the 2020 cumulative incidence of COVID-19 and the ln-transformed NHL incidence in 1974-2003, weighing by population size and adjusting by socioeconomic deprivation and other covariates.

**Results:** The cumulative incidence of COVID-19 increased in relation to past incidence of NHL (*p* < 0.001), socioeconomic deprivation (*p* = 0.006), and proportion of elderly residents (*p* < 0.001) and decreased with urban residency (*p* = 0.001). Several sensitivity analyses confirmed the finding of an association between COVID-19 and NHL.

**Conclusion:** This ecological study found an ecological association between NHL and COVID-19. If further investigation would confirm our findings, shared susceptibility factors should be investigated among the plausible underlying mechanisms.

**Strengths and limitations of this study:**

- This study exploited the availability of incidence data for non-Hodgkin’s lymphoma over a 30-year time frame and the 2020 cumulative incidence data for COVID-19 available for all administrative units in the Sardinia region, Italy.
- Using an ecological study design, we observed that COVID-19 occurrence increased with past incidence of non-Hodgkin’s lymphoma.
- Limitations of the ecological study design require further investigation for confirmation and for identification of susceptibility factors possibly shared between the two diseases.

## Introduction

The worldwide geographical variation in the incidence of and mortality from the 2019 coronavirus disease (COVID-19) and the inter-individual susceptibility to its more severe forms suggests a potential interplay of genetic susceptibility and environmental factors [1]. Ecological research on the contributing factors has detected associations with population density and vehicular traffic [2], temperature and humidity [3], and air pollution [1,4]. Also, during the first epidemic wave but not afterwards, the vaccination against seasonal influenza provided protection [2,5]. On the other hand, genetic variants in the 2’-5’-oligoadenylate synthetase (OAS1) gene [3], the ACE (Angiotensin-Converting Enzyme) receptors [6], the inflammatory cytokine cascade [7,8], and the Janus kinase (JAK/STAT) pathway of complement activation [9] have been suggested to confer greater susceptibility to SARS-CoV-2 infection. In Brazilian couples with only one individual affected despite sharing the same bed, HLA-DRB1 alleles were associated with symptomatic infection, while HLA-A alleles were more prevalent in seronegative women [10]. Also, the cytokine storm in COVID-19 notably features a sharp increase in interleukin 10 (IL-10) [7] and TNF-*α* [8] serum levels, which would depend on the level of expression of the respective genes. Furthermore, COVID-19 has shown a direct link with TNF-*α* and TNF-*β* gene polymorphisms [11] and with the rs1800896 IL-10 single gene polymorphisms (SNP), but not the rs1800871 and rs1800872 IL-10 SNPs [12,13].

Interestingly, cell surface antigens and cytokine gene polymorphisms play a role in the aetiopathogenesis of non-Hodgkin’s lymphoma (NHL) as well. For instance, the risk of various lymphoma subtypes increases in association with HLA class I and class II variants [14-16] and polymorphisms in genes expressing IL-10 and TNF [17] and in genes related to the JAK-STAT signalling pathway [18]. Consistently with the apparently similar genetic susceptibility features, numerous studies have confirmed a strong link between previous viral and microbial infections, including HIV infection, and NHL risk, whether through immunosuppression or other mechanisms [19,20]. On the other side, a few reports have described an increase in COVID-19 incidence and severity among patients with a diagnosis of primary cutaneous lymphoma (PCL) [21,22] and haematological disease in general [23,24], but not among patients suffering from any cancer, including lymphoma patients [25]. However, NHL subtypes may differ in terms of vulnerability to SARS-CoV-2 infection. Treatment-induced immunosuppression [26,27] and a low-rate seroconversion [28] in onco-haematological patients would only partly explain such differences.

A recently published Bayesian analysis showed that, over the three decades between 1974 and 2003, the NHL incidence tended to cluster in the north-eastern part of the Italian region of Sardinia and its major urban centre, with the low incidence areas located in the south [29]. The ecological analysis did not identify any plausible explanatory factor among the known conditions associated with an increased NHL risk. If common susceptibility factors contributed to linking COVID-19 and NHL, one would expect the two diseases to show consistent geographic patterns. Taking profit of the availability of the 2020 cumulative incidence data for COVID-19 by administrative unit, we explored the correlation between past incidence of NHL and incidence of COVID-19 over the administrative units within Sardinia.

## Methods

The age- and gender-standardized incidence rates of NHL between 1974-2003 were available for all the 356 autonomous administrative units (communes) in Sardinia as of 1974 [29]. Briefly, due to the lack of a regional Cancer Registry, the chief haematologist of the Cagliari Oncology Hospital, with the support of the health authorities and the clinical departments of the whole region, created a database of all incident haematological cancers, diagnosed among the Sardinian population of both genders and any age from 1974-2003 [30]. Its completeness was validated through the comparison with mortality and hospitalization data [31] and, limited to the last decade and the northern part of the region, with Cancer Registry records [32]. For each commune, we abstracted data on the cumulative incidence of COVID-19 from the first diagnosis to 31 December 2020, before the start of the vaccination campaign, from a publicly available report [33]. Only crude cumulative incidence of COVID-19 was locally available. Also, as information was unavailable at the commune level, we could not analyse COVID-19 mortality, or the rate of positives over the total tests performed or the disaggregated figures by symptomatic or asymptomatic positive cases. The communes’ population varied from 77 to 195,500, with 196 (55%) inhabited by less than 2,000 people and 26 (7%) comprising 56% of the resident population. Despite being the most prevalent group of onco-haematological neoplasms, NHL is relatively rare, with a global incidence of 7.8 per 100,000 per year (all ages) [34]. For this reason, the precision of the estimate of the local NHL incidence rate was very low for many communes with small population size. For instance, 25 communes (all with less than 2,000 inhabitants) had no incident NHL cases in 1974-2003, while the incidence varied from 1.2 to 17.4 per 100,000 in the 74 communes where the cases were only 1 or 2. The 2020 incidence of COVID-19 manifested the same zero-case occurrence in four communes with less than 500 inhabitants. We graphically addressed this problem using bubble plots, with the bubble radius proportional to the commune’s population size, and analytically by conducting a weighted regression using the ratio between the local and the regional population as the weight.

We applied natural log transformation (ln) to both the age- and sex-standardised NHL incidence rates (independent variable) and the COVID-19 incidence rates (outcome), as the residuals of the regression model using the raw data violated normality at the lower and upper tails. We added a small constant to each value of both variables to allow the ln transformation even for zero values. The regression models also included the proportion of inhabitants aged ≥75 years, the male/female ratio among the residents, the Italian Institute of Statistics (ISTAT) deprivation index (http://istat.it), the distance from the nearest hospital, and the urban/rural type of commune. The urban or rural type covariate for each commune was defined based on the presence/absence of five community services (administrative, educational, health, judicial, and religious) that would attract daily commuters from the surrounding area. In a sensitivity analysis, we used conventional (instead of a weighted) regression analysis after grouping the communes with less than 10,000 residents into 36 larger units corresponding to historical sub-regional areas and used these and the remaining 26 larger communes in the analysis. This analysis included 61 units as one of the 26 larger communes was part of an area consisting only of two communes. For the aggregated areas, the deprivation index, the proportion of elderly, and the male/female ratio were calculated as weighted averages within each area. Further sensitivity analyses included 1) considering only the communes with more than 2,000 residents and 2) a separate analysis limited to each approximate quartile of the resident population.

The best fitting model was selected using a stepwise backward approach, with the goodness-of-fit of the models assessed using the R^2^ value, which indicates the proportion of the variability in the COVID-19 incidence rate explained by the covariates in the model. The analysis was conducted using SPSS® version 20.0.

A STROBE statement on ecological studies, such as the present study, is still missing. However, we acknowledged the limitations of such a study design and complied with the requirements for ecological studies. In our analysis we only accessed aggregated data, which use for the purposes of scientific publication was approved by the Ethics Committee of the University Hospital of Cagliari (protocol N. PG 2019/18070, 18 December 2019).

### Patient and public involvement

None.

## Results

The 1974-2003 NHL age- and gender-adjusted incidence rate among the Sardinian adult population (aged 25 years or more) was 13.4 per 100,000 (95% CI 13.0 – 13.8) [29], ranging from 0 to 27.9 across the 356 communes. The 2020 cumulative incidence of COVID-19 was 2,117 per 100,000 population, ranging from 0 in four communes with 77, 148, 224, and 506 residents, respectively, to 11,935 in a commune with 1,240 residents. Figure 1 shows the bubble plots of the COVID-19 incidence against NHL incidence. The regression line in Figure 1A describes the weighted regression equation in all the communes, and that in Figure 1B describes the unweighted regression equation in the 61 units (25 large communes and 36 aggregated areas); both show an upward trend of increasing 2020 COVID-19 incidence with increasing past NHL incidence.

**Figure 1.**
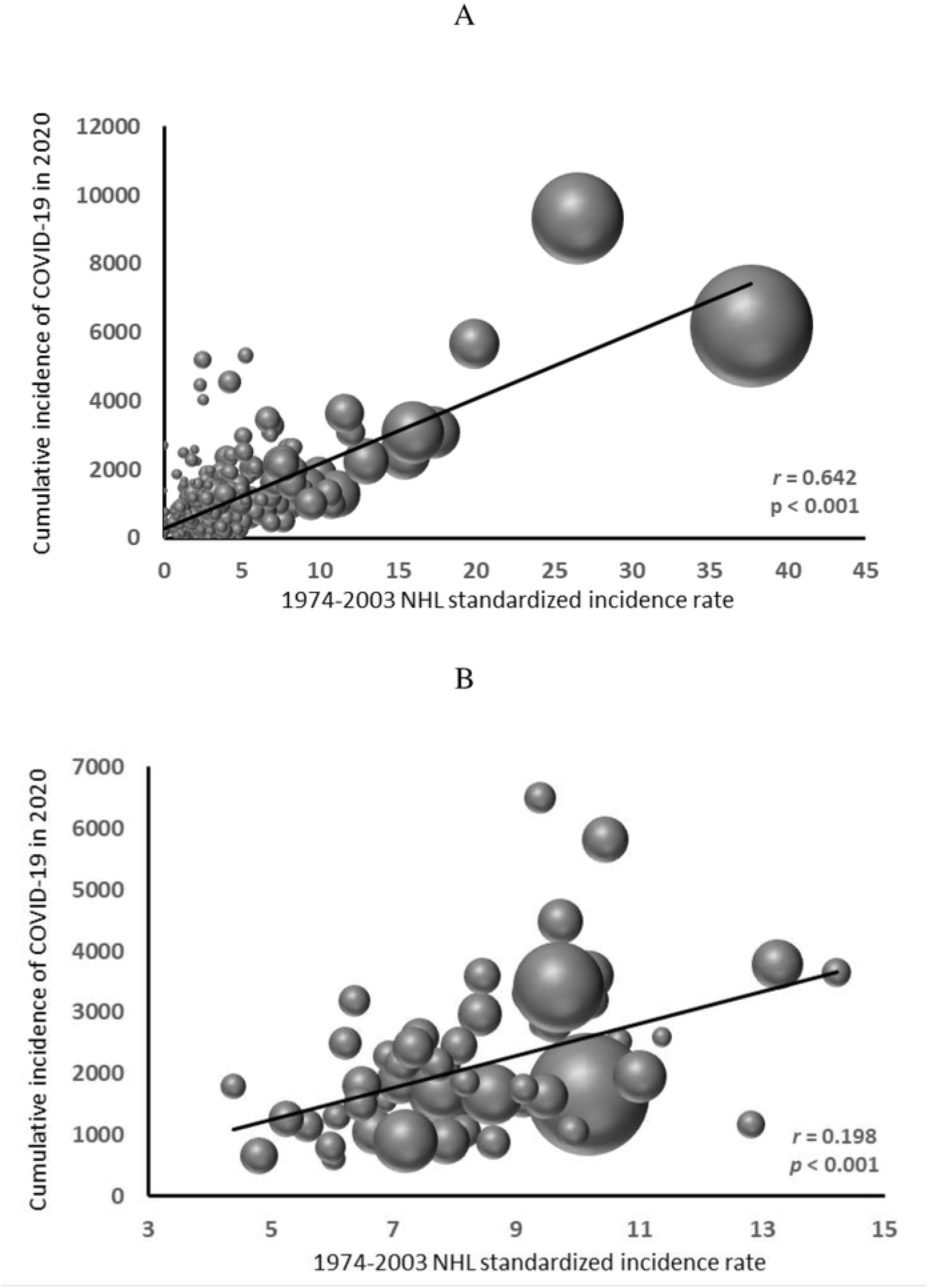
Bubble graph of 2020 cumulative incidence of COVID-19 in relation to 1974-200 incidence of non-Hodgkin’s lymphoma in Sardinia, Italy. A. all communes (N = 356), weighed (weigh= resident population); B. 61 residential units, including 25 communes with 10000 residents or more and 36 sub-regional areas incorporating the remaining 331 communes.

The best fitted multiple regression model (adjusted R^2^ = 0.31) included the ln-transformed NHL incidence rate (*β* = 1.021, *p* < 0.001), the proportion of elderly among the resident population (*β* = 0.427, *p* < 0.001), the type of residence (*β* = -0.331 for urban *vs* rural residence, *p* = 0.008), and the deprivation index (*β* = 0.134, *p* = 0.035) as the covariates (Table 1, Model 1). The distance to the nearest hospital and the male/female ratio were not associated with COVID-19 incidence, nor did their inclusion in the model result in a better fit. The analyses of the 61 administrative units (Table 1, Model 2) and by gender (data not shown) confirmed the results.

**Table 1.** Parameters of the multiple regression analysis with cumulative incidence of COVID-19 as the outcome: model 1: weighted regression, all communes; outcome: ln-transformed COVID-19 incidence; covariates: ln-transformed standardized NHL incidence, deprivation index, proportion of elderly residents, and urban vs rural residence. The last three are not ln-transformed; model 2: conventional regression, 61 administrative units: outcome: cumulative incidence of COVID-19; covariates: standardized NHL incidence, deprivation index, proportion of elderly residents.

| *Independent covariates* | *Model 1* |  |  | *Model 2* |  |  |
| --- | --- | --- | --- | --- | --- | --- |
| | $\beta$ | *se* | *p* | $\beta$ | *se* | *p* |
| Ln-transformed NHL incidence | 1.021 | 0.196 | < 0.001 | 244.6 | 69.2 | < 0.001 |
| Deprivation index | 0.134 | 0.063 | 0.035 | 93.8 | 116.4 | 0.424 |
| Proportion of elderly residents | 0.427 | 0.101 | < 0.001 | 288.0 | 120.5 | 0.020 |
| Urban vs rural residence | -0.331 | 0.124 | 0.008 | - |  |  |
| Adjusted $R^2$ | | 0.30 | | | 0.25 | |

Further sensitivity analyses, by dropping the small size communes (<2,000) and by quartile of population size, consistently showed an increase in COVID-19 in relation to past NHL incidence (data not shown).

## Discussion

Our results suggest a significant geographic correlation between the cumulative incidence of COVID-19 in 2020 and the 1974-2003 NHL incidence within the region of Sardinia. We limited the analysis to the first year of the COVID-19 pandemic, before vaccines became available, to avoid the effect of the geographical and age-related variation in compliance with the vaccination. The results were consistent in the sensitivity analyses that used different approaches to overcome the analytical problem created by the sparse data in the numerous small-size communes. Based on the weighted regression analysis, a 1% increase in the past NHL incidence would have yielded a 1.02% increase in the incident cases of COVID-19 per 100,000 residents in 2020, corresponding to 22 additional cases. Amongst the other variables included in our model, consistently with previous reports, the proportion of elderly residents and socio-economic deprivation also showed an association with an increasing cumulative incidence of COVID-19 [1,35]. On the other hand, sex and distance from the nearest hospital did not show an association, and urban residency was inversely related to COVID-19 incidence. However, this last finding was not confirmed in the sensitivity analyses.

Only the total number of incident COVID-19 cases was publicly accessible at the local level, which did not allow a proper age and gender standardization of the rates. This drawback limits the interpretation of our results; to mitigate the resulting bias, we used the proportion of elderly and the male/female ratio among the residents as adjusting covariates in the multiple regression models. While the male/female ratio did not show an association with COVID-19 incidence nor contributed to reducing the model variance, the proportion of elderly did. As incidence increased nationwide among the elderly but varied little by gender (4.7% of the male population vs 4.6% of the female population) [36], we presumed to have at least partially rectified the drawback. A further limitation results from the unavailability at the commune level of the number of swab tests, which would have allowed using a perhaps more reliable measure of incidence. Mortality data and disaggregated figures of COVID-19 incident cases by the presence or absence of symptoms were also unavailable; therefore, we could not investigate the most severe forms of COVID-19.

The suspicion of bias due to the “ecological fallacy” has repeatedly been raised for findings in ecological studies, as associations at the population level might not translate into associations at the individual level [37]. However, ecological studies are helpful in the preliminary exploration of the link between new health emergencies, such as the global COVID-19 pandemic, and possible pre-existing conditions and risk factors in a population. Such studies can provide aetiological clues to direct further research aimed at informing public health preventive strategies [38]. Therefore, while recommending further epidemiological investigations into the association between NHL and COVID-19, we caution against an over-interpretation of our findings.

Should further research confirm our findings, a shared susceptibility between NHL and COVID-19 might be postulated, including genetic susceptibility. If so, polymorphisms in genes implicated in the synthesis of cytokines, HLA cell surface antigens, and others facilitating the virus entry into human cells in impairing the immune response might play a role in the pathogenesis of both diseases. Consequently, the greater susceptibility of lymphoma patients to SARS-CoV-2 infection and the most severe forms of COVID-19 might be due not only to the immunosuppressive effects of the therapy protocols but also to a common susceptible background for NHL and COVID-19.

## Data Availability

The data underlying the results presented in the study are available from Prof. Pierluigi Cocco (contact).

## Contributorship Statement

**Sara De Matteis:** Conceptualization, Validation, Project administration, Roles/Writing - original draft, Writing - review & editing. **Cosetta Minelli:** Methodology, Formal analysis, Writing - review & editing. **Giorgio Broccia:** Data curation, Writing - review & editing. **Paolo Vineis:** Writing - review & editing. **Pierluigi Cocco:** Conceptualization, Supervision, Formal analysis, Roles/Writing - original draft, Writing - review & editing.

## Data sharing statement

Data are preserved in the archives of the department of Medical Sciences and Public Health of the Cagliari University in aggregated form, and they are publicly available as such. Please contact Prof. Sara De Matteis for any request.

## Ethical approval statement

The Ethics Committee of the Cagliari University Hospital approved the study protocol (Protocol No. PG 2019/18070, 18 December 2019) in agreement with the Code of Ethics of the World Medical Association (Declaration of Helsinki).

The authors are grateful to Mr Maurizio Addis and Mr Franco Porcu for local advice and support in accessing the official COVID-19 incidence data.

